# Prediction of SARS-CoV-2 transmission dynamics based on population-level cycle threshold values: A Machine Learning and mechanistic modeling study

**DOI:** 10.1101/2023.03.06.23286837

**Authors:** Afraz A. Khan, Hind Sbihi, Michael A. Irvine, Agatha N. Jassem, Yayuk Joffres, Braeden Klaver, Naveed Janjua, Aamir Bharmal, Carmen H. Ng, Amanda Wilmer, John Galbraith, Marc G. Romney, Bonnie Henry, Linda M. N. Hoang, Mel Krajden, Catherine A. Hogan

## Abstract

**Background:** Polymerase chain reaction (PCR) cycle threshold (Ct) values can be used to estimate the viral burden of Severe Acute Respiratory Syndrome Coronavirus type 2 (SARS-CoV-2) and predict population-level epidemic trends. We investigated the use of machine learning (ML) and epidemic transmission modeling based on Ct value distribution for SARS-CoV-2 incidence prediction during an Omicron-predominant period.

**Methods:** Using simulated data, we developed a ML model to predict the reproductive number based on Ct value distribution, and validated it on out-of-sample province-level data. We also developed an epidemiological model and fitted it to province-level data to accurately predict incidence.

**Results:** Based on simulated data, the ML model predicted the reproductive number with highest performance on out-of-sample province-level data. The epidemiological model was validated on outbreak data, and fitted to province-level data, and accurately predicted incidence.

Conclusions

These modeling approaches can complement traditional surveillance, especially when diagnostic testing practices change over time. The models can be tailored to different epidemiological settings and used in real time to guide public health interventions.

**Funding:** This work was supported by funding from Genome BC, Michael Smith Foundation for Health Research and British Columbia Centre for Disease Control Foundation to C.A.H. This work was also funded by the Public Health Agency of Canada COVID-19 Immunity Task Force COVID-19 Hot Spots Competition Grant (2021-HQ-000120) to M.G.R.

## Introduction

SARS-CoV-2 viral burden can be quantitated by the use of polymerase chain reaction (PCR) cycle threshold (Ct) values, which are inversely proportional to the amount of target viral sequence present in the patient sample. Although this information is frequently available from routine molecular methods for the diagnosis of SARS-CoV-2 infection, clinical results are almost universally reported qualitatively as present or absent due to sources of sampling variability, lack of inter-test standardization, insufficient supporting clinical correlation data, and lack of regulatory approval for purposes other than qualitative reporting, all of which limit interpretation of Ct values for clinical care. Though the use of Ct values to guide individual-level management is not currently routinely recommended (1, 2), the assessment of aggregated Ct values at a population level may be useful to help assess early epidemiological transmission trends to improve epidemic forecasting (3–5), and parallels the concept of measuring community viral load used for other viruses (6–8). Accurate projection of epidemic trends is critical to effectively plan public health efforts including healthcare resource allocation. Indeed, an epidemic in the growth phase is more likely to be associated with high viral load burden at a population level; conversely, the decline phase of an epidemic is likely to demonstrate lower viral burden. A modeling approach was previously published to inform epidemic SARS-CoV-2 trajectory based on aggregated Ct value data (3), and supported the usefulness of population-level Ct value analysis. However, SARS-CoV-2 testing practices globally have evolved substantially during the pandemic, most frequently by restricting testing to symptomatic individuals, which limits the usefulness of modeling approaches that rely on stable population sampling strategies. Starting in December 2021 in British Columbia (BC), use of PCR testing was partially restricted in the context of roll-out of rapid antigen tests, limiting understanding of population trends. New tools are needed to estimate incidence in a manner that is independent of the biases associated with testing guidance. This includes modeling approaches robust to varying testing guidelines, sample selection strategies and epidemiologic settings, and that account for other variables that impact viral burden such as variant of concern (VoC) and vaccination status. The main variants of concern (VoC) described to date have been associated with varying impact on viral burden, with the Delta and certain Omicron subvariants associated with highest viral load (9–14). Furthermore, evidence suggests that SARS-CoV-2 vaccination is associated with a reduction in viral burden, and correspondingly higher Ct values and potentially lower transmission risk, in individuals who develop post-vaccination infection (14–19). In this study, we investigated two modeling approaches based on Ct value distribution of asymptomatic individuals, machine learning and epidemic transmission modeling, to predict SARS-CoV-2 incidence based on province-wide data and an outbreak in a long-term care facility in British Columbia, Canada. We assessed the novel application of five machine learning models (Lasso, LGBM, XGBoost, CatBoost, RF), and validated two previously-described epidemic models (SEIR) (3), to determine the highest performing models across a range of epidemiological settings to predict SARS-CoV-2 incidence.

## Methods

### Study design

Individuals with polymerase chain reaction (PCR)-confirmed SARS-CoV-2 infection by nasopharyngeal swab or saline gargle between November 19^th^ 2021 and January 8th 2022 were included, capturing emergence of Omicron wave in the province. Descriptive analyses of Ct value distribution included the two main specimen type categories: nasopharyngeal swabs and saline gargles, while modeling analyses focused on nasopharyngeal swabs given the higher diagnostic yield and collection standardization. Analysis was based on province-wide data incorporating all SARS-CoV-2 diagnostic tests based on the *E* gene target performed in BC. Three pandemic phases in BC were considered based on vaccination roll-out and VoC distribution (**Supplemental Table 1)**. To capture the largest representation of asymptomatic individuals in BC throughout the pandemic, the study focused on phase 3. These individuals were tested in the context of occupational screening or pre-travel. The study population sampled thus represented a heterogeneous mix of vaccinated and unvaccinated individuals, and predominantly Omicron (BA.1) variant (**Figures 1A and 1B**).

**Figure 1.**
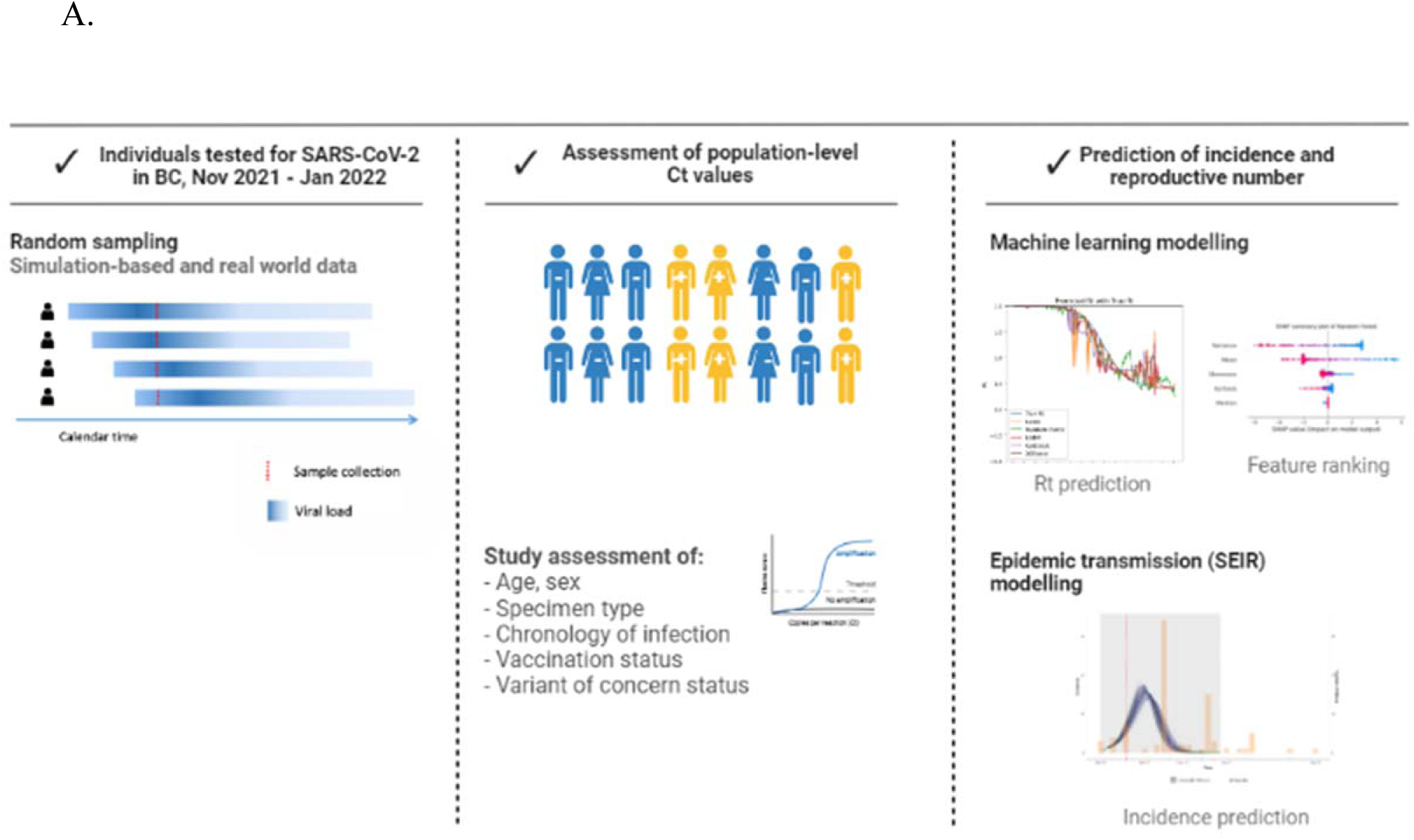
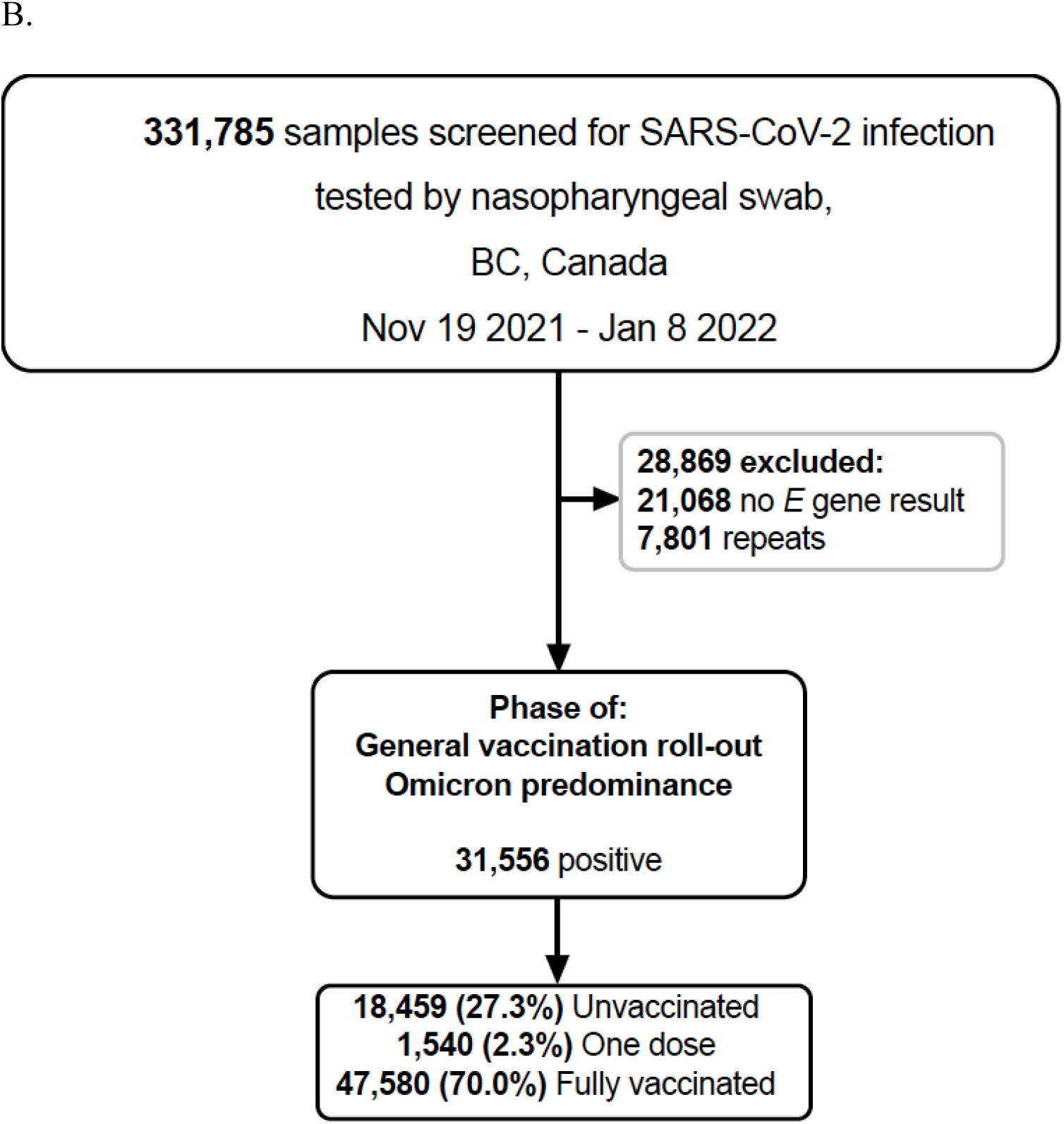
Overall design (A) and flowchart (B) of the study. BC: British Columbia; E gene: Envelope gene; SARS-CoV-2: Nov: November; Rt: reproductive number; SARS-CoV-2: severe acute respiratory syndrome coronavirus type 2

Given the importance of population composition in informing model selection, the selected models were chosen based on several factors including sampling type and frequency, sample size, and computational complexity. Current models which make use of cross-sectional Ct values to infer epidemic trajectories (3) rely on random sampling of the population to accurately predict epidemic trends. However, in the context of symptom-based testing, the distribution of Ct values is not a complete representation of the infected population. Thus, this study was performed on a population of asymptomatic individuals as they served as the best proxy for frequent non-symptom-based sampling.

### Testing practices and public health measures

COVID-19 diagnostic testing and laboratory test guidelines changed in BC over the course of the pandemic, and can be summarized as follows: 1) exposure-based testing (onset of the pandemic), 2) targeted testing (starting March 16, 2020), 3) expanded testing (starting April 9, 2020), 4) symptom-based testing (starting April 21, 2020), 5) revised symptom-based testing (starting December 17, 2020), and 6) High risk/targeted population testing (increased risk of severe disease or work in a high-risk setting) (starting January 18, 2021) (20, 21). Thus, asymptomatic and mildly symptomatic testing was initiated starting in December 2021 with the organized roll-out of rapid antigen tests, which corresponds with the current study. While COVID-19 testing was initially centralized at the BCCDC Public Health Laboratory (PHL), testing capacity and data capture reflects results from all provincial testing laboratories which generate *E* gene Ct values.

### Laboratory data - SARS-CoV-2 diagnostic testing

SARS-CoV-2 diagnostic testing was performed in laboratories throughout all five health authorities in BC, and only assays based on the *E* gene target were included for this study. The testing strategy and test result interpretation criteria used for the participating laboratories are described separately (**Supplemental Table 2**), and included the BCCDC PHL laboratory-developed test (LDT) (22), LightMix SarbecoV E-gene plus EAV control assay (TIB Molbiol, Berlin, Germany), Alinity m SARS-CoV-2 (Abbott, Chicago, IL), BD SARS-CoV-2 (Becton, Dickinson and Co., Franklin Lakes, NJ), cobas 6800 and 8800 (Roche Diagnostics, Basel Switzerland), GeneXpert Xpress SARS-CoV-2 (Cepheid, Sunnyvale, CA), Panther Fusion (Hologic, San Diego, CA) and Allplex SARS-CoV-2 (Seegene, Seoul, South Korea). For individuals having undergone repeat SARS-CoV-2 testing within a one-week period, only the first positive test per person was included.

### Laboratory data - Variant of Concern identification

The BCCDC Public Health Laboratory (PHL) continuously monitors for variants of concern (VOCs), variants of interest (VOIs), and variants under monitoring (VUMs). Various approaches were used over time including VoC screening and confirmation by whole genome sequencing (WGS) when applicable at the BCCDC PHL as previously described (22). Testing strategy is optimized based on available capacity and clinical and public health needs, and changed over the course of the SARS-CoV-2 pandemic. One such strategy included deployment of in brief, a subset of samples in the earlier phase of the epidemic (January 2021 to May 2021) was tested by targeted single nucleotide polymorphism (SNP) quantitative polymerase chain reaction (qPCR) for VoC screening, followed by confirmation by WGS. From June 2021 onward, sample VoC status was detected by WGS alone. From September 2021, owing to increased case burden and limited capacity, there was a transition from WGS of all samples to a subset positive SARS-CoV-2 samples. This subset comprised of targeted surveillance (cases from outbreaks, vaccine escape, reinfection and travel-related), and representative baseline surveillance. In addition, 100% of positive samples underwent WGS in the first week of each month. Starting November 15 2021 in the context of the Omicron variant emergence, WGS was resumed for all samples. Owing to the high transmissibility of Omicron and the surge in case load, starting December 21 2021, there was transition from full sequencing to sequencing a subset of representative positive samples in addition to priority cases (including outbreaks, long-term care, vaccine escape, travel-related, hospitalization)). Full VoC characterization for the province of BC is described separately (**Supplemental Figure 2**).

### Vaccination status

Vaccination status was defined based on the date of vaccine receipt relative to the date of the sample collection included for the study (**Supplemental Figure 3**) (23). For primary dose series all mRNA (Pfizer, Moderna) and viral vector vaccines (AstraZeneca, Janssen) were considered. For the Janssen vaccine only, fully vaccinated status was defined as having received one dose 14 days or more prior to sample collection. For all other vaccines, **Unvaccinated status** was defined as having received no SARS-CoV-2 vaccine, or having received a SARS-CoV-2 vaccine less than 21 days prior to the sample collection date. **Partially vaccinated** status was defined as having received the SARS-CoV-2 vaccine dose 1 greater or equal to 21 days prior to sample collection, but having received dose 2 less than 14 days prior to the sample collection. **Fully vaccinated** status was defined as greater or equal to 14 days since the receipt of dose 2, but having received dose 3 less than 14 days prior to the sample collection. Cross-over vaccination was considered in the same category as homologous vaccine schedules.

### Outbreak case study

To further validate the models, a separate analysis was performed using a well-characterized outbreak in a long-term care facility that occurred in BC. This outbreak was selected on the basis of time of occurrence of pre-vaccination roll-out to the general population, large size and generalizability of the affected population. This outbreak included large-scale asymptomatic testing. Testing was done weekly until no additional cases were identified within 14 days of the last exposure. There were 7 rounds of weekly testing at the outbreak facility, all negative residents and staff were tested for each round. Anyone who developed symptoms was also tested. The epidemiologic data and curve describing the outbreak are presented separately (**Supplemental Figure 4**). As for the main study, analysis was based on SARS-CoV-2 diagnostic tests based on the *E* gene target. However, due to missing data in the long-term care facility data, wherever the *E* gene target was unavailable the *ORF1* gene target was used instead.

### Data sources

Two main data sources were employed for this study: 1) the Provincial Health Laboratory Viewer and Reporter (PLOVER) database which includes the laboratory diagnostic datasets, and 2) the Provincial Immunization Registry (PIR) dataset which includes vaccination data. The laboratory datasets house data on SARS-CoV-2 testing (including date of collection, specimen type, diagnostic quantitative PCR gene target results, VoC screening, and SARS-CoV-2 lineage based on WGS), and individual-level epidemiological data (including age, sex, patient as well as ordering physician health authority). Gene target results include Ct values of the *E* and ORF1 targets, and the internal control RNAseP. The PIR dataset includes individual-level vaccination data (including number, type, series, dose and date of each vaccine received). Both of these datasets form the basis of the covariates which inform the ML models in the study. For the outbreak case study, additional data were directly gathered from public health partners (**Supplemental Figure 4**) as these were not otherwise available through provincial datasets. Data linkages were performed between the laboratory and PIR datasets through a sequential deterministic linkage based on a minimum of three personal identifiers (personal health number, last name with first three digits of first name, and date of birth). These linkages were performed prospectively on a weekly basis, and specimens with unsuccessful linkages were excluded from the study.

### Data & Code Availability

The genomic sequencing data are publicly available in GISAID under the submitter British Columbia Center for Disease Control Public Health Laboratory (BCCDC PHL). The individual level demographic and epidemiological data can be made accessible following the data governance and data access policy guidelines (http://www.bccdc.ca/about/accountability/data-access-requests). Code for this study is available (https://github.com/Afraz496/Vital-E-paper).

### Ethics

This research was approved by University of British Columbia Research Ethics (H20-0297 BCC19C-COVID-19 Research).

### Models

#### Machine learning and epidemic transmission models

This study compared two different approaches for inference, machine learning (ML) and epidemic transmission modeling, both of which were used to predict the reproductive number (R_t_) to estimate incidence. The first modeling approach was based on a collection of ML approaches, including Lasso (24), Random Forest (RF), Light Gradient Boosting Modeling (LGBM), eXtreme Gradient Boosted Modeling (XGBM) and CatBoost. Due to the testing guidelines which were tailored for mainly symptomatic individuals, and given the absence of sufficiently-large random samples, analysis was pursued with simulated data instead. Ct data were generated to simulate a sufficiently large random sample of a population using the virosolver package (R software, version 4.1.2). This was applied on varying sample sizes (100, 1000 and 10000) on a simulated population of 50,000 individuals. The simulation horizon was set at 140 calendar days to encompass a typical single COVID-19 wave. These data were used to create summary statistics of the Ct distribution including mean, median, variance, skewness and kurtosis. The trained data was generated from a unique simulation file with a fixed random seed and three distinct sample sizes, so three models were investigated in this study. Hyperparameter tuning was performed via a grid search of hyperparameters on each model (**Supplemental Table 5**). The best performing model was chosen by finding the optimal set of hyperparameters for which the Mean squared error (MSE) between the true simulated Rt and Predicted simulated Rt was minimized. SHapley Additive exPlanation (SHAP) analysis was performed for feature ranking and importance (25). The second modeling approach was adapted from an existing methodology (3), and is based on a single epidemic model. The compartmental SEIR model captures different stages in individual infections (namely **S**usceptible, **E**xposed, **I**nfectious and **R**ecovered). The SEIR model was validated on a patient outbreak facility in BC where point prevalence testing was done in infrequent intervals. The SEIR model was then fitted to provincial data from asymptomatic individuals. Modifications to the viral kinetics for the SEIR model were applied to these provincial data to account for the specific nature of the Omicron (BA.1) variant.

## Results

### Cohort description

During the study period, a total of 331,785 SARS-CoV-2 tests were performed in BC, of which 79,443 were positive. Restricting these to the first positive test per person, there were 71,642 included in the study (**Figures 1A and 1B**). Of these, 35,369 were nasopharyngeal specimens and 36,108 were saline gargle specimens (**Table 1 and** **Figure 2**). The cohort was predominantly composed of adults aged 18-59 years (72.2%), followed by adults aged 60 years and above (12.1%), and children 0-17 years (15.7%). Over half the cases resided in two of the five health authorities accounting for 35.9% and 30.9%, respectively. The Omicron (BA.1) variant predominated throughout the study (**Table 1 and Supplemental Figure 2**). By the end of the study period, a total of 18,459 (27.3%) were unvaccinated, 1,540 (2.3%) had received 1 dose of vaccine, and 47,580 (70%) were fully vaccinated.

**Figure 2.**
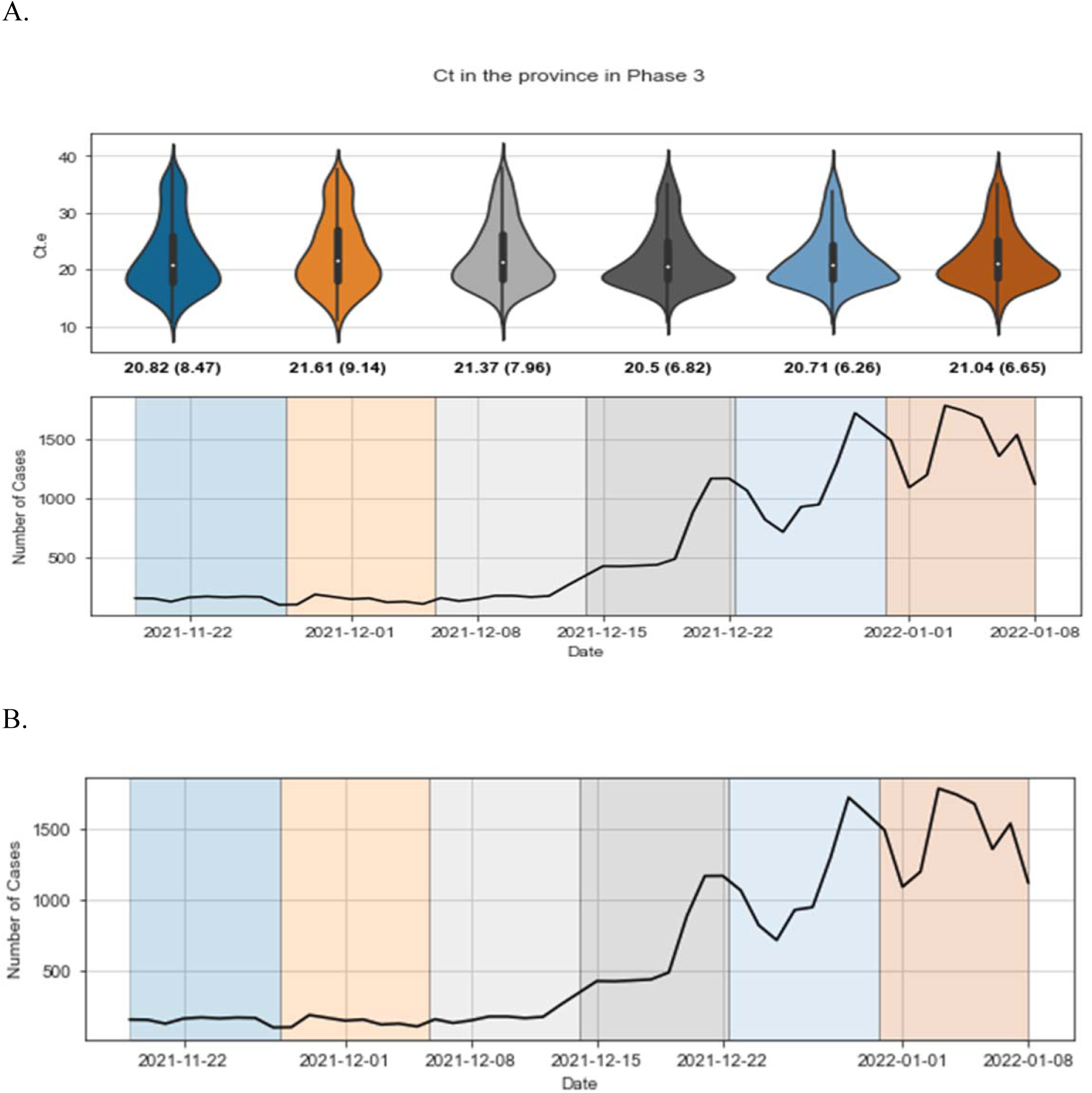
Violin plots demonstrating the cycle threshold value distribution (A) and absolute number of cases of confirmed SARS-CoV-2 infection (B) in British Columbia across different time points of the study period. Ct. e: Envelope (*E*) gene cycle threshold value; SARS-CoV-2: SARS-CoV-2: severe acute respiratory syndrome coronavirus type 2

**Table 1.** Epidemiological, clinical and laboratory data of the cohort of asymptomatic individuals tested during the test period of the study.

| Group | Subgroup | Phase 3 |
| --- | --- | --- |
| Testing | First positives | 71642 |
|  | Negatives | 252342 |
|  | Repeats | 7801 |
| Specimen type | NP | 35369 |
|  | SG | 36108 |
|  | Other | 165 |
| No *E* gene result |  | 21068 |
| Age (years) | 0-4 | 1963 |
|  | 5-18 | 9161 |
|  | 19-39 | 31914 |
|  | 40-59 | 19864 |
|  | 60-79 | 7453 |
|  | 80+ | 1279 |
| Sex | Male | 33415 |
|  | Female | 37653 |
|  | Unknown | 574 |
| Patient health authority | 1 | 31841 |
|  | 2 | 8951 |
|  | 3 | 3635 |
|  | 4 | 17202 |
|  | 5 | 9752 |
|  | Unknown | 261 |
| Vaccination status** | Unvaccinated | 18459 |
|  | 1 dose | 1540 |
|  | Fully vaccinated | 47580 |
|  | Unknown | 4063 |
| VoC lineage | Alpha | 0 |
|  | Beta | 0 |
|  | Delta | 9049 |
|  | Gamma | 0 |
|  | Omicron (BA.1) | 12945 |
|  | Unknown | 28580 |

\*For all group variables except testing, data presented as first positive result per person
\*\*Does not include individuals who received $\geq 3$ doses of vaccine
NP: nasopharyngeal; SG: saline gargle; VoC: variant of concern

**Table 2.** SEIR and ML model comparisons for SARS-CoV-2 incidence prediction

| Model |  | SEIR | ML |
| --- | --- | --- | --- |
| Sampling type |  | Random sampling |  |
| Number of COVID-positive samples |  | Small (~30) | Large (>1000) |
| Sampling frequency |  | Single/multiple snapshots | Daily snapshots |
| Flexibility: | i) Modeling of Transmission | i) Fixed in time | i) Time-independent |
|  | ii) Ability to add in multiple predictors | ii) Unable to incorporate multiple predictors | ii) Allows incorporation of various predictors and is flexible in their representation |
| Scalability |  | Single outbreak setting | Population level |
| Computational Complexity* |  | Low | Low-moderate |
| Predictive power requirements |  | Good in single setting with well-mixed population and stable contact behavior/infection control | No requirements other than sufficient sample size for Ct summary statistics by snapshot |
| Additional Sampling Requirements |  | None | Ordered in time, restricted to fixed interval sampling |

\* Relative computational complexity based on assumed sample size provided in scalability row
COVID-19: coronavirus disease 2019; SEIR: susceptible-exposed-infected-recovered; ML: machine learning; Ct: cycle threshold

#### First modeling approach: machine learning

The fitted ML models were applied to out-of-sample Ct data from the simulated Ct values (**Figure 3**). With increasing sample sizes, the MSE across each model reduced by 82% showing an increased ability in higher moments (mean, median, variance, skewness and kurtosis) of the Ct distribution to predict epidemic trends. Random Forest showed the largest improvement in MSE performance while demonstrating lowest performance in smaller sample size. Besides the smallest sample size, all models generally perform similarly across an increased sample size (**Figure 3**). For the largest sample size, apart from Lasso all other models have a much tighter IQR and smaller MSE median score (at around 0.03). Across all sample sizes, the variance of the Ct distribution was the top ranking feature (**Supplemental Figure 5**).

**Figure 3.**
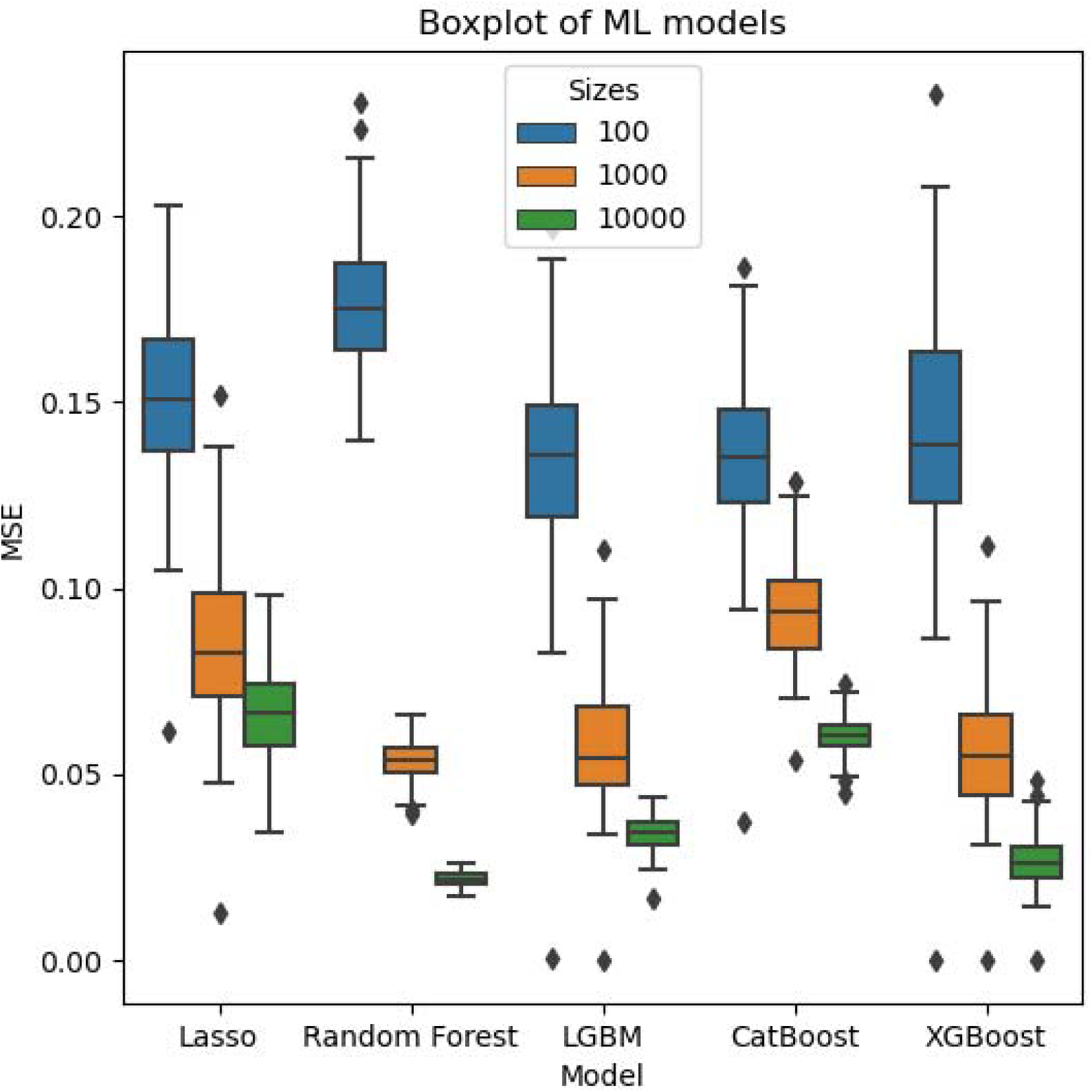
Boxplot representation of MSE scores across models on out-of-sample simulated cycle threshold data. LGBM: Light Gradient Boosting Model; MSE: Mean squared error; XGBM: eXtreme Gradient Boosting Model, ML: Machine Learning

#### Second modeling approach: epidemic transmission models

##### SEIR

The most precise results were observed with sampling from a total of five horizons. The model posteriors indicated an incidence peak from December 27 2021 to January 1 2022, which overlapped with the observed peak of reported cases in the province (**Figure 4**). Similarly, the exponential growth phase coincided with the increase in reported cases from our cohort from December 20 2021 to December 27 2021, and the decline of the incidence coincided with the decline in cohort cases from January 1 2022 to January 5 2022. The posterior predictive Ct distribution also closely matched the observed Ct distribution on each of the time horizons, supporting accurate incidence projection independent of biases of testing guidance.

**Figure 4.**
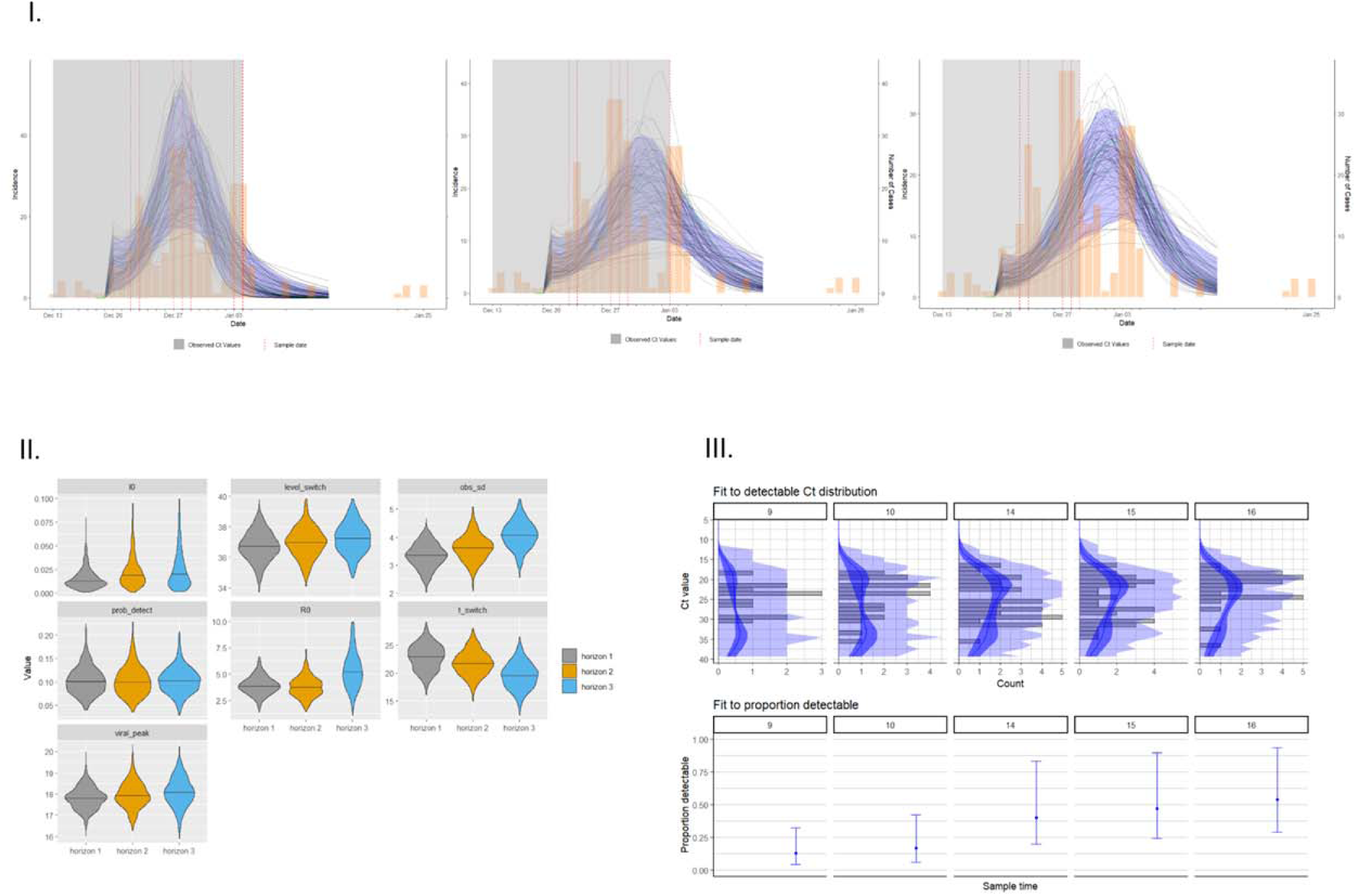
Overall population modeling findings. A multiple-cross section SEIR model was fitted to the overall population-level data (I), and showed an incidence peak from December 27 2021 to January 1 2022, which overlapped with the observed peak of reported cases in the province. The Monte Carlo chain model-predicted incidence curve is represented (black lines), and was overlaid with the reported number of confirmed SARS-CoV-2-positive (yellow bars) cases. Violin plots of the viral kinetic parameters for the SEIR model are presented (II). Three unique time horizons were chosen, each of which is depicted by a different color. The MCMC approach searches over the viral kinetic parameters presented above, and is based on prior values described separately (**Supplemental Table 6**). To align with the described Omicron viral kinetics, the incubation period was fixed and set at three days, and the infectious viral kinetic parameter was fixed. An upper bound of I_0_ was set at 0.100. The fit to detectable cycle threshold distribution and the fit to proportion variable are presented over different time points (A). Ct: cycle threshold; SARS-CoV-2: severe acute respiratory syndrome coronavirus type 2; SEIR: susceptible-exposed-infected-recovered

##### Outbreak case study

This outbreak occurred in a long-term care facility, and resulted in a total of 156 individuals (93 residents and 63 staff) infected with SARS-CoV-2 (**Supplemental Figure 4**). Of these individuals, 58.1% of infections were asymptomatic in the residents, whereas 9.5% were asymptomatic within the staff. There were 26 (28.0%) deaths in the residents group, and no deaths among the staff. A multiple cross-section SEIR model was fitted to the outbreak data, and showed a peak in incidence on the 12th day of the outbreak which preceded by two days the observed peak at the outbreak facility (**Figure 5**). The real incidence fell within the 95% credible interval of the predicted MCMC chains of the SEIR model. The model also accurately predicted the decline in cases by the 20th day of the outbreak (**Figure 5**).

**Figure 5.**
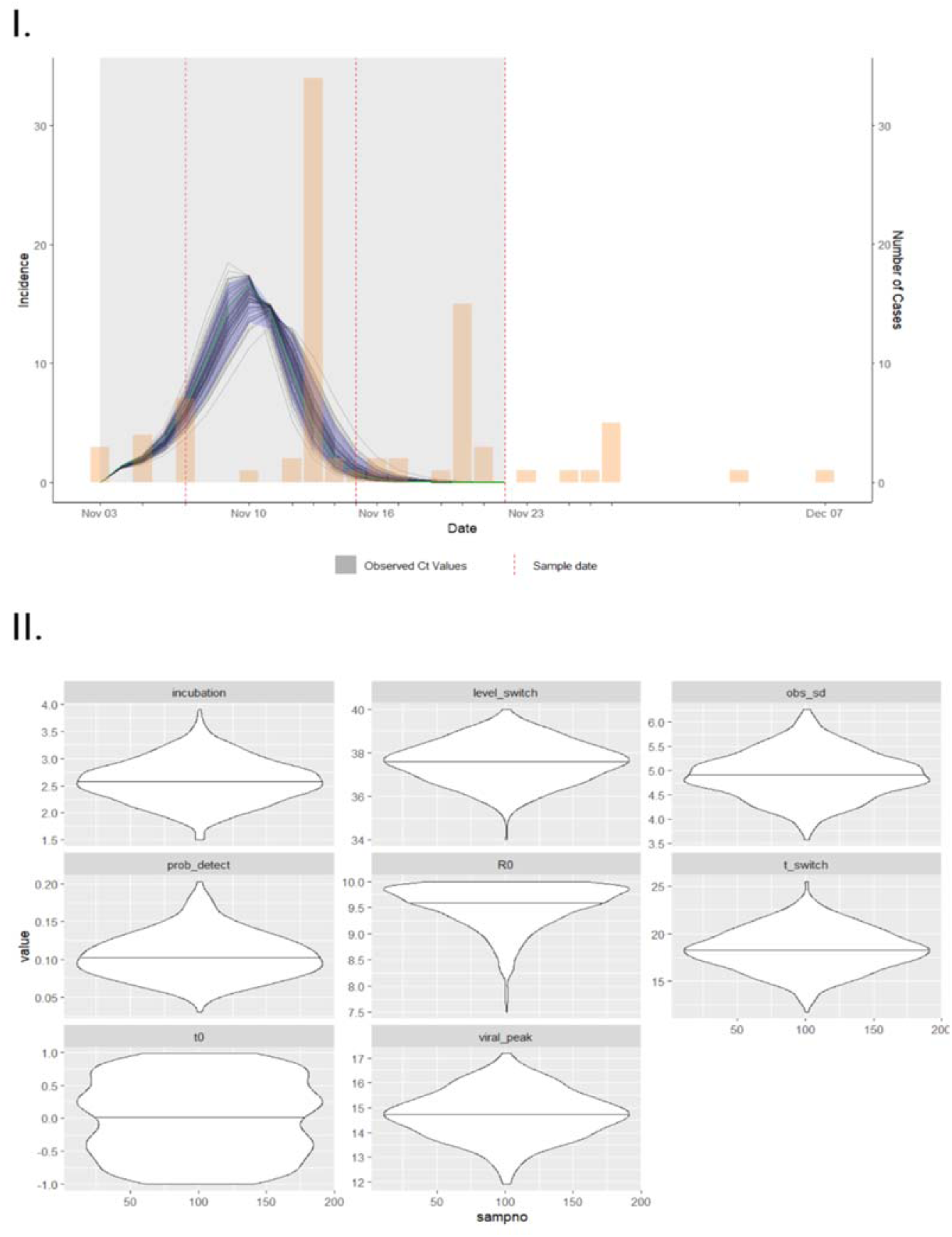
Long-term care facility outbreak investigation modeling findings. a multiple-cross section SEIR model was fitted to the outbreak data (I), and showed a peak in incidence on the 12th day of the outbreak which preceded by two days the observed peak at the outbreak facility. The population included in this outbreak investigation was sampled at three pre-determined time points (dashed red lines). The Monte Carlo chain model-predicted incidence curve is represented by black lines, and was overlaid with the reported number of confirmed SARS-CoV-2-positive cases in this outbreak setting (yellow bars). Violin plots of the viral kinetic parameters for the SEIR model are also presented in the outbreak case study (II). Ct: cycle threshold; SARS-CoV-2: severe acute respiratory syndrome coronavirus type 2; SEIR: susceptible-exposed-infected-recovered

## Discussion

In this study, we demonstrated the utility of two distinct modeling approaches based on aggregated cycle threshold values, machine learning and epidemic transmission modeling, to predict epidemic trends across varying sampled patient populations, random, and targeted and non-random testing. Based on out-of-sample mean squared error (MSE) change in the reproduction number between the true and predicted values, the ML model performed best on randomly-sampled province-level data. Within epidemic transmission models, the SEIR model performed highest with randomly-sampled outbreak data. Taken together, these approaches accurately predicted epidemic transmission dynamics at the outbreak case study level, and at a provincial level for the province of BC, Canada. Early in the pandemic, SARS-CoV-2 diagnostic molecular testing was more largely based on random sampling which, despite possible underascertainment due to lack in testing access, could be used to estimate full case counts to monitor and predict transmission dynamics. As testing needs overwhelmed laboratory capacity with increasing case burden and the emergence of variants of concern, molecular testing practice recommendations shifted to testing individuals who were symptomatic and/or with a minimal illness severity, resulting in sampling of a selected population. These changes in testing indications, foremost predicated on symptom-based testing, led to substantially more limited capacity to assess case counts for epidemic monitoring, generating a critical unmet need for other approaches to infer epidemic trends to support clinical and public health planning.

This study comprehensively investigated varying sampling types and modeling approaches, drawing on both previously-published work and description of the novel application of machine learning modeling for SARS-CoV-2 transmission dynamics prediction. Our work identified that diagnostic testing indication, sampling type, and the individual population tested are critical factors, and that model selection must be tailored to the epidemiological circumstances of testing. More specifically, random vs targeted or non-random sampling must be accounted for to ensure appropriate model selection, as SEIR modeling was only suitable for random sampling. For example, performance of the SEIR model in this study was robust across sample sizes and the long-term care facility dataset. Results from this modeling approach demonstrated a slight difference in incidence peak timing and amplitude. This is likely explained due to the lag time between onset of the infectious period and reporting given that site-wide facility testing was performed at set time periods rather than on a daily basis, and represents a pragmatic approach to real-world settings. Thus, this approach is well suited for long-term care or assisted living or community-living facility outbreak investigations such as shelters, or within small hospital systems. In contrast, the novel application of machine learning approaches described in this study performed the best with large datasets (>1,000 COVID-19 positive cases), making this the approach of choice for large population settings such as at the province, state or large hospital network system level. Indeed, machine learning models can offer greater flexibility by incorporating different summary statistics and other data as features, fully harnessing the potential of larger datasets.

Importantly, all approaches described in this study could predict future trends within a one to four-week timeframe, demonstrating utility for timely prediction of SARS-CoV-2 transmission dynamics that could be harnessed to help inform future outbreak resource allocation and decision-making. Thus, use of these models can be used to support critical decision-making across several settings, including hospitals, long-term care facilities, public health departments and others, to help inform planning of resource allocation, vaccination efforts, and isolation practices. More specifically, this approach lays the groundwork for a sentinel surveillance monitoring strategy that could be automated and alert appropriate authorities at pre-determined signals of predicted incidence changes, and may be expanded to other infections for which testing is widespread and predictive tools are needed.

This study focused on a time period of Omicron (BA.1) predominance, and revealed that despite its shorter incubation period compared to other variants of concern, the Ct distribution of this variant could successfully be described through an SEIR compartmental model and machine learning approaches. Furthermore, in the context of a sampled population with heterogeneous vaccination status, the current study demonstrated accurate prediction of incidence based on overall Ct distribution and viral kinetics without incorporating individual-level vaccination status. Further work is necessary to study the impact of vaccination status on accuracy of incidence prediction.

The main strength of this study is that it provides a comprehensive modeling toolkit that can be leveraged across population and sampling settings, and that may incorporate covariates such as variant of concern and vaccination status. This approach could predict transmission dynamics in a way that could not be performed through case count analysis from biased sampling as was occurring in the province of BC. This modeling is also advantageous as it can be performed in real-time, rather than rely on monitoring of clinical indicators of severity such as hospitalization and intensive care unit admission which considerably lag behind true incidence rise. A limitation of previous studies is the use of a single or limited methodology for analysis that may perform well in a specific setting such as long-term care facilities, but lacked flexibility and predictive performance for generalizability to larger settings and in the context of changing testing practices (3). Our body of work filled this gap and further presented a methodology to incorporate assessment of variant of concern and vaccination status, two important potential confounders on Ct value distribution, although these characteristics were noted to be less important than the moments of infection (mean, median, variance, skewness and kurtosis of the aggregated Ct distribution). Additional strengths of this study also include the independent assessment of the models in a long-term care facility outbreak to validate the previously-published models (3). Furthermore, the main analysis drew on a provincial dataset linking laboratory data and vaccination status in real-time, thus leveraging the design for the highest possible public health uptake and impact. Taken together, this approach lays the framework for expansion to use for other pathogens for which surveillance needs are critical including other respiratory pathogens and monkeypox. Indeed, further work may also build on this approach and further integrate complementary datasets including wastewater Ct distribution to further enhance prediction ability.

However, there are several limitations. Firstly, the methodology is based on the assumption of random or random sampling which is challenging to confirm. Indeed, testing practices were modified following clinical and public health guidance of the province, and may have led to bias in sampling. Restriction of the study population to the asymptomatic subgroup consisting of travelers and occupational health testing led to greater confidence in the employed sampling strategy tested and the validity of this assumption. The need for random sampling remains a limitation for broader uptake of this approach, though it may be more attainable in the context of outbreak investigation where full populations are sampled at once. Nonetheless, even when a full population is sampled there may be specific population-level characteristics that need to be accounted for. One such limitation in the current work is that although the long-term care environment provides more a consistent testing environment, it tends to be a highly vaccinated population which may introduce bias. Finally, this study aggregated Ct-level data across multiple laboratories and assays, which may not adequately capture intra- and inter-assay variation.

In summary, this study proposes a comprehensive suite of modeling strategies based on population-level Ct values to accurately predict SARS-CoV-2 transmission dynamics across epidemiological settings. These modeling approaches can be used in real time to guide clinical and public health interventions. Such tools are needed to estimate incidence in a manner that is independent of the biases associated with testing guidance, and to complement traditional surveillance based on case numbers or clinical indicators. Further work will be needed to expand validation of the machine learning models based on larger datasets and different settings with newly-emerging variants, to assess real-time predictive power for direct clinical and public health impact.

## Funding

This work was supported by funding by Genome BC, Michael Smith Foundation for Health Research and British Columbia Centre for Disease Control Foundation to C.A.H. This work was also funded by the Public Health Agency of Canada *COVID-19 Immunity Task Force COVID-19 Hot Spots Competition Grant (2021-HQ-000120)* to M.G.R.

## Data Availability

The genomic sequencing data is publicly available in GISAID under the submitter British Columbia Center for Disease Control Public Health Laboratory (BCCDC PHL). The individual level demographic and epidemiological data can be made accessible following the data governance and data access policy guidelines (http://www.bccdc.ca/about/accountability/data-access-requests). Code used for study models will be made available upon request to the corresponding author.

## Acknowledgments

We thank the laboratory teams (virology, bacteriology and molecular) at the BCCDC Public Health Laboratory for their contribution toward testing, on which this research is based. We also thank the data analytics team for supporting the data infrastructure and review that enabled this work. Finally, we thank the British Columbia Association of Medical Microbiologists for testing and sharing samples and data that enabled province-wide data collection, and public health partners throughout the province for their dedicated effort to outbreak management and infection control, and for sharing outbreak-level data that supported this research.

---

**Supplemental Table 1.** Vaccination phase definitions used for the study

| **SARS-CoV-2 phase** | **Vaccination Phase\*** | **Wave\*\*** | **Target population** |
| --- | --- | --- | --- |
| Phase 1 | Phase 1 | Waves 1, 2, 3<br>Dec 14 2020<br>- Mar 7 2021 | Residents, staff and essential visitors to long-term care settings; individuals assessed and awaiting a long-term care placement; health care workers providing care for COVID-19 patients; and remote and isolated Indigenous communities. |
| | Phase 2 | 3<br>Mar 8 2021 -<br>Apr 2021 | Individuals age $\geq 80$; Indigenous peoples age $\geq 65$ and Indigenous Elders; Indigenous communities; hospital staff, community general practitioners and medical specialists; vulnerable populations in select congregate settings; and staff in community home support and nursing services for seniors. |
|  | Phase 3 | 3<br>Apr 15 2021 -<br>May 10 2021 | Individuals aged 60-79 years, Indigenous peoples aged 18-64 and people aged 16-74 who are clinically extremely vulnerable. |
| Phase 2.1 | Phase 4 | Waves 3, 4<br>May 11<br>2021- Jul 17<br>2021 | Everyone aged $\geq 12$ years-old. From September 2021, third vaccine dose available for people who are clinically extremely vulnerable |
| Phase 2.2 | Phase 4 | 4<br>Jul 18 2021 -<br>Nov 18 2021 |  |
| Phase 3* | Phase 5 | 4<br>Nov 19 2021<br>- Jan 8 2022 | Everyone aged $\geq 5$ years-old. From the end of November 2021, children aged 5-11 are eligible for vaccination. Everyone aged $\geq 18$ and invited to get a 'booster' (third vaccine dose) within 6-8 months after receipt of their second dose. |

\*The current study included Phase 3 only
\*\*Vaccination phases were defined by vaccine eligibility of the target populations in BC, and are detailed separately (21)
SARS-CoV-2: SARS-CoV-2: severe acute respiratory syndrome coronavirus type 2

**Supplemental Table 2.**
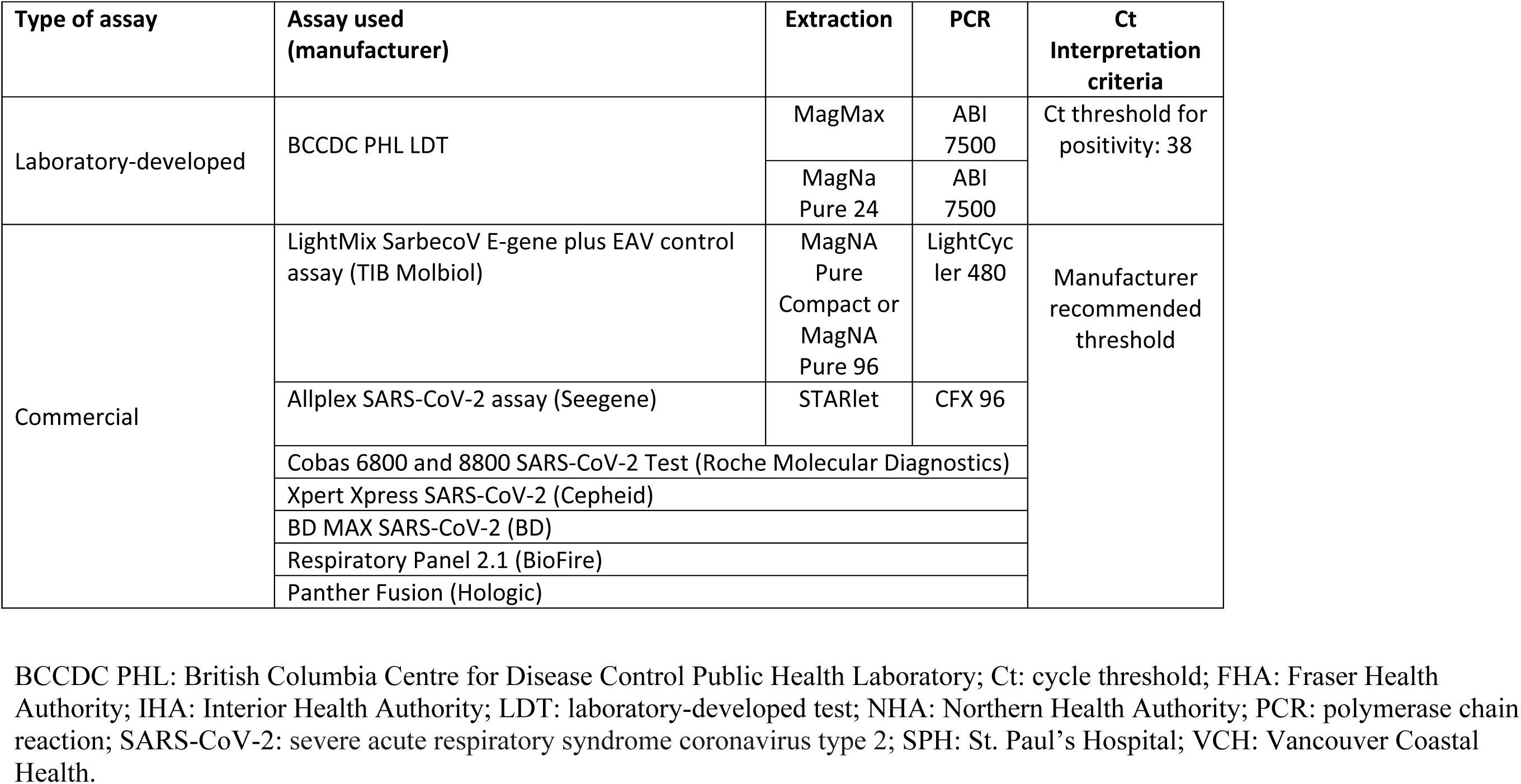
SARS-CoV-2 diagnostic testing strategy based on the envelope (*E*) gene target and test result interpretation criteria used for the participating laboratories

| Type of assay | Assay used (manufacturer) | Extraction | PCR | Ct Interpretation criteria |
| --- | --- | --- | --- | --- |
| Laboratory-developed | BCCDC PHL LDT | MagMax | ABI 7500 | Ct threshold for positivity: 38 |
|  |  | MagNa Pure 24 | ABI 7500 |  |
| Commercial | LightMix SarbecoV E-gene plus EAV control assay (TIB Molbiol) | MagNA Pure Compact or MagNA Pure 96 | LightCycler 480 | Manufacturer recommended threshold |
|  | Allplex SARS-CoV-2 assay (Seegene) | STARlet | CFX 96 |  |
|  | Cobas 6800 and 8800 SARS-CoV-2 Test (Roche Molecular Diagnostics) |  |  |  |
|  | Xpert Xpress SARS-CoV-2 (Cepheid) |  |  |  |
|  | BD MAX SARS-CoV-2 (BD) |  |  |  |
|  | Respiratory Panel 2.1 (BioFire) |  |  |  |
|  | Panther Fusion (Hologic) |  |  |  |

BCCDC PHL: British Columbia Centre for Disease Control Public Health Laboratory; Ct: cycle threshold; FHA: Fraser Health Authority; IHA: Interior Health Authority; LDT: laboratory-developed test; NHA: Northern Health Authority; PCR: polymerase chain reaction; SARS-CoV-2: severe acute respiratory syndrome coronavirus type 2; SPH: St. Paul’s Hospital; VCH: Vancouver Coastal Health.

**Supplemental Table 3.**
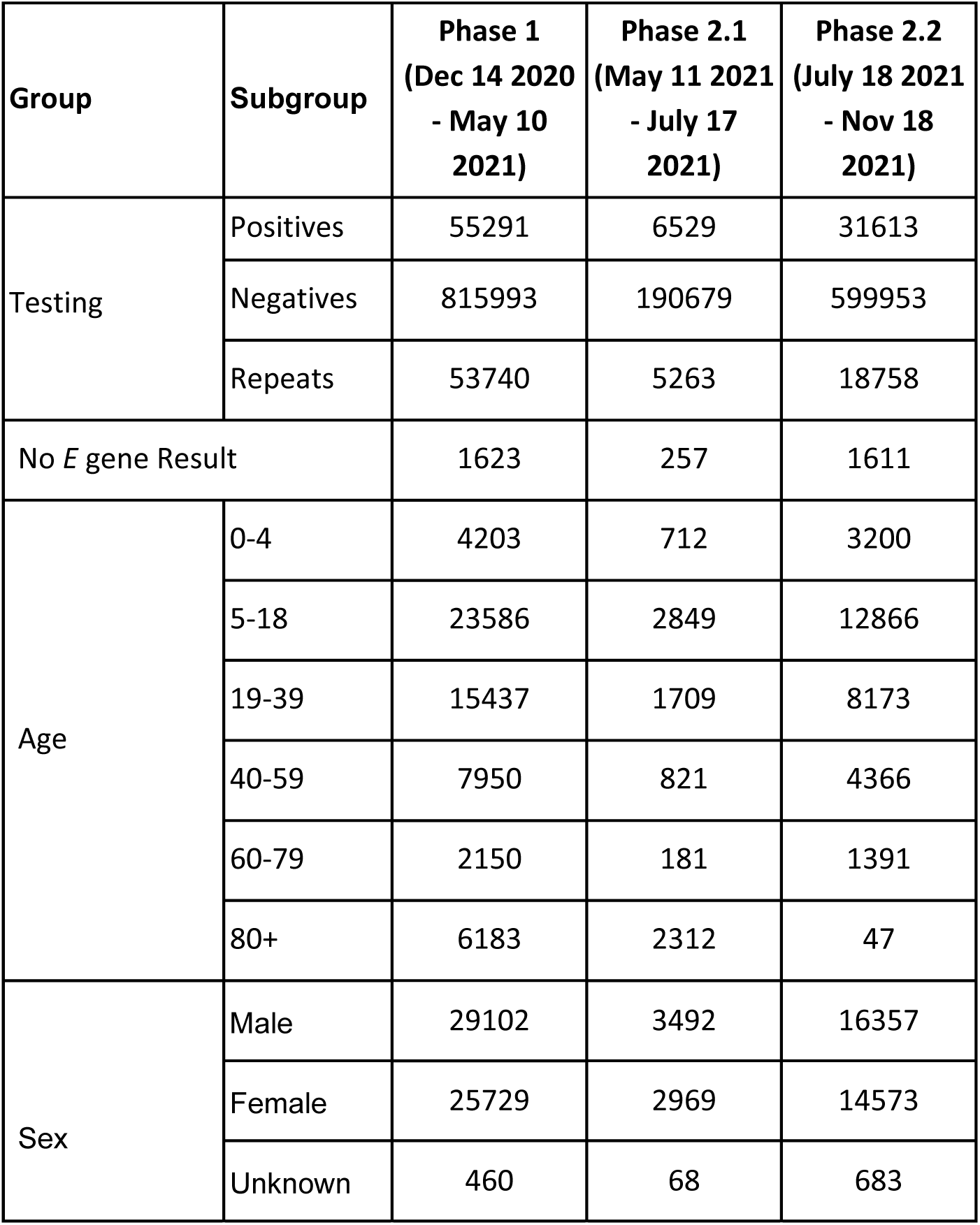
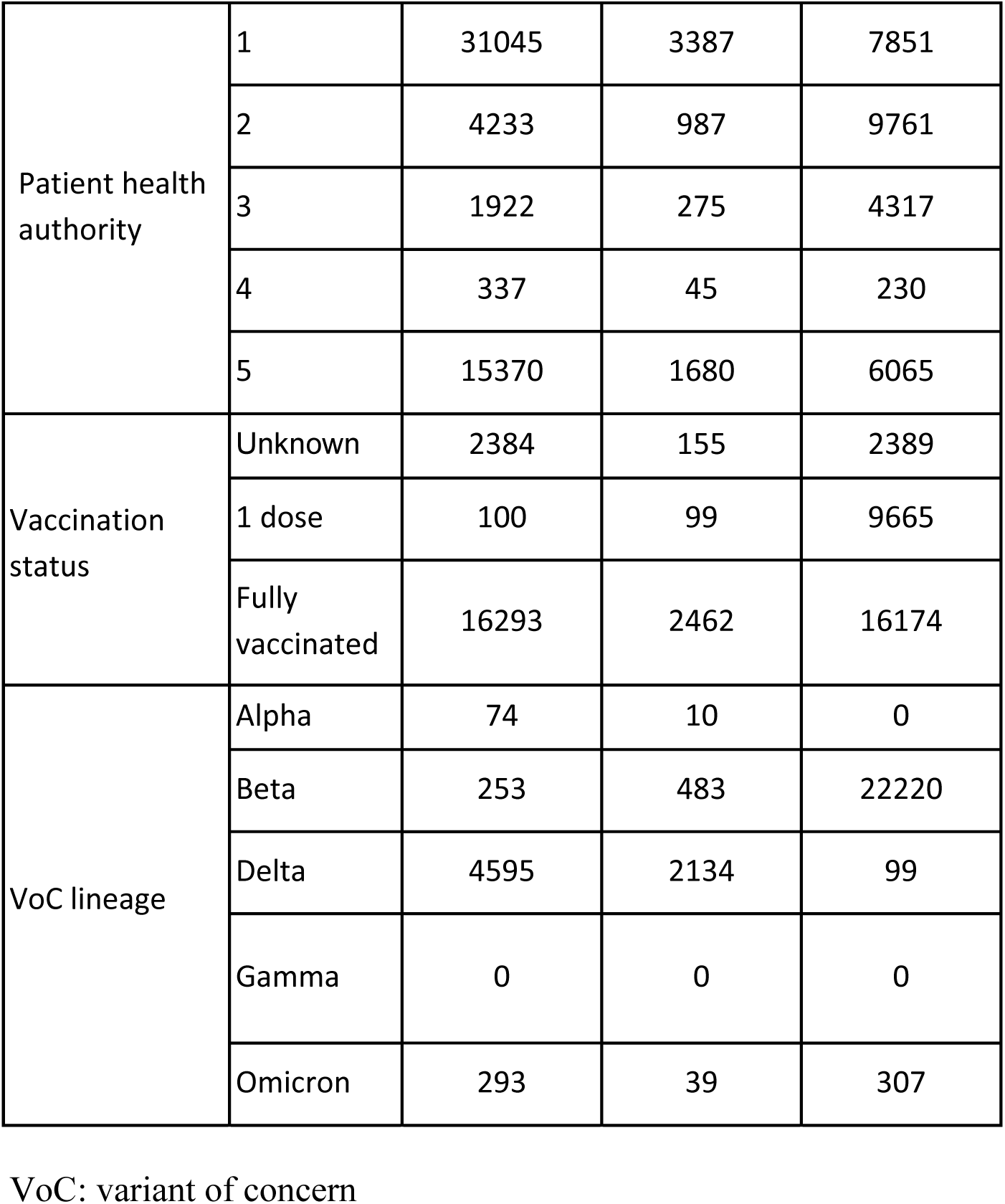
Epidemiological, clinical and laboratory data of the earlier British Columbia SARS-CoV-2 pandemic phases

| Group | Subgroup | Phase 1<br>(Dec 14 2020<br>- May 10<br>2021) | Phase 2.1<br>(May 11 2021<br>- July 17<br>2021) | Phase 2.2<br>(July 18 2021<br>- Nov 18<br>2021) |
| --- | --- | --- | --- | --- |
| Testing | Positives | 55291 | 6529 | 31613 |
|  | Negatives | 815993 | 190679 | 599953 |
|  | Repeats | 53740 | 5263 | 18758 |
| No *E* gene Result |  | 1623 | 257 | 1611 |
| Age | 0-4 | 4203 | 712 | 3200 |
|  | 5-18 | 23586 | 2849 | 12866 |
|  | 19-39 | 15437 | 1709 | 8173 |
|  | 40-59 | 7950 | 821 | 4366 |
|  | 60-79 | 2150 | 181 | 1391 |
|  | 80+ | 6183 | 2312 | 47 |
| Sex | Male | 29102 | 3492 | 16357 |
|  | Female | 25729 | 2969 | 14573 |
|  | Unknown | 460 | 68 | 683 |
| Patient health authority | 1 | 31045 | 3387 | 7851 |
|  | 2 | 4233 | 987 | 9761 |
|  | 3 | 1922 | 275 | 4317 |
|  | 4 | 337 | 45 | 230 |
|  | 5 | 15370 | 1680 | 6065 |
| Vaccination status | Unknown | 2384 | 155 | 2389 |
|  | 1 dose | 100 | 99 | 9665 |
|  | Fully vaccinated | 16293 | 2462 | 16174 |
| VoC lineage | Alpha | 74 | 10 | 0 |
|  | Beta | 253 | 483 | 22220 |
|  | Delta | 4595 | 2134 | 99 |
|  | Gamma | 0 | 0 | 0 |
|  | Omicron | 293 | 39 | 307 |

VoC: variant of concern

**Supplemental Table 5.** Hyperparameter selection

| Hyperparameter | Lasso | RF | LGBM | XGBoost | Catboost |
| --- | --- | --- | --- | --- | --- |
| Alpha | [0.001,0.01,0.1] | - | - | - | - |
| Max Depth | - | [2,4,8,16,32] | [2,4,8] | [6,10] | - |
| Number of Estimators | - | [4,16,64,256] | - | - | - |
| Minimum Sample Split | - | [2,4,8,16,32] | - | - | - |
| Number of Leaves | - | - | [4,8,16,32,64,128] | - | - |
| Minimum Data in Leaf | - | - | [2,4,8,16,32] | - | - |
| Depth | - | - | - | - | [1,5,10] |
| Iterations | [500,1000,2000] | - | - | - | [250,500,1000] |
| Learning Rate | - | - | - | - | [0,001, 0.01, 0.1] |
| L2 Regularization | - | - | - | - | [1,5,10] |
| Min child weight | - | - | - | [1,3] | - |

RF: Random Forest; LGBM: Light Gradient Boosting Model; XGBM: eXtreme Gradient
Boosting Model

**Supplemental Table 6.** Control table of priors for SEIR model

| Values | Names* | Fixed | Lower bound | Upper bound | Steps | Lower start | Upper start |
| --- | --- | --- | --- | --- | --- | --- | --- |
| 0 | tshift | 1 | 0 | 3 | 0.1 | 0 | 10 |
| 5 | desired_mode | 1 | 0 | 7 | 0.1 | 0 | 10 |
| 19.73 | viral_peak | 0 | 0 | 40 | 0.1 | 15 | 25 |
| 5 | obs_sd | 0 | 0 | 25 | 0.1 | 1 | 10 |
| 0.79 | sd_mod | 1 | 0 | 1 | 0.1 | 0.4 | 0.6 |
| 14 | sd_mod_wane | 1 | 0 | 14 | 0.1 | 0 | 14 |
| 40 | true_0 | 1 | 40 | 100 | 0.1 | 40 | 100 |
| 40 | intercept | 1 | 35 | 100 | 0.1 | 35 | 100 |
| 3 | LOD | 1 | 0 | 10 | 0.1 | 0 | 10 |
| 5 | incu | 1 | 0 | 10 | 0.1 | 0 | 10 |
| 13.29 | t_switch | 0 | 0 | 30 | 0.1 | 10 | 30 |
| 38 | level_switch | 0 | 0 | 40 | 0.1 | 33 | 40 |
| 1000 | wane_rate2 | 1 | 0 | 10000 | 0.1 | 10 | 50 |
| 0.103 | prob_detect | 0 | 0 | 1 | 0.1 | 0.01 | 0.1 |
| 1 | t_unit | 1 | 0 | 1 | 0.1 | 0 | 1 |
| 2 | R0 | 0 | 1 | 10 | 0.1 | 1.5 | 3 |
| 4 | infectious | 1 | 0 | 25 | 0.1 | 5 | 10 |
| 3 | incubation | 1 | 0 | 25 | 0.1 | 5 | 10 |
| 1 | t0 | 1 | 0 | 100 | 0.1 | 0 | 50 |
| 0.0001 | I0 | 0 | 0 | 0.1 | 0.1 | 0 | 0.001 |

\*Adapted parameters (3)
SEIR: susceptible-exposed-infected-recovered

**Supplemental Figure 1.**
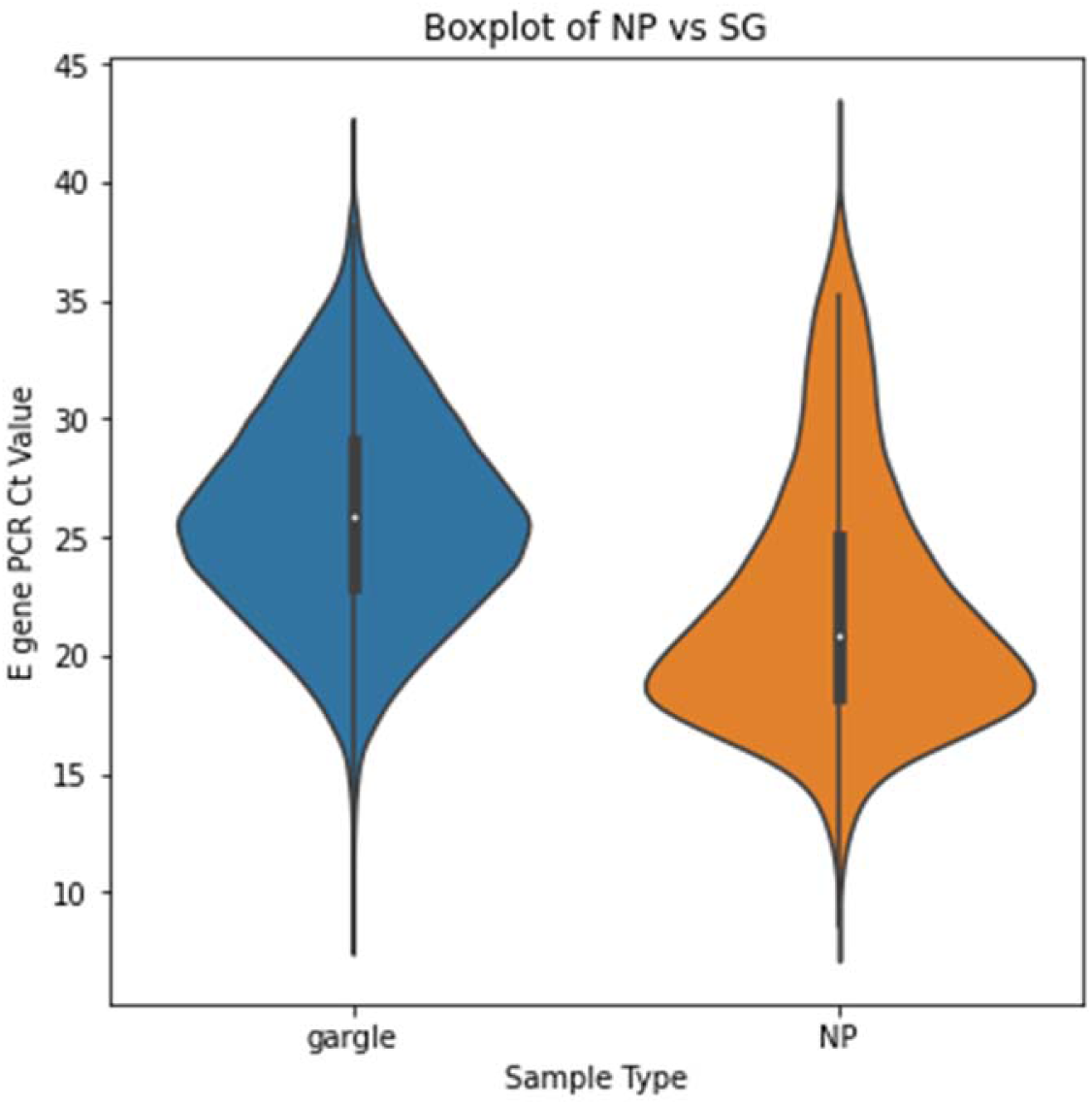
Violin plot demonstrating the overall cycle threshold value distribution for saline gargle compared to nasopharyngeal specimens for the entire study period. Ct. e: Envelope (*E*) gene cycle threshold value; NP: nasopharyngeal

**Supplemental Figure 2.**
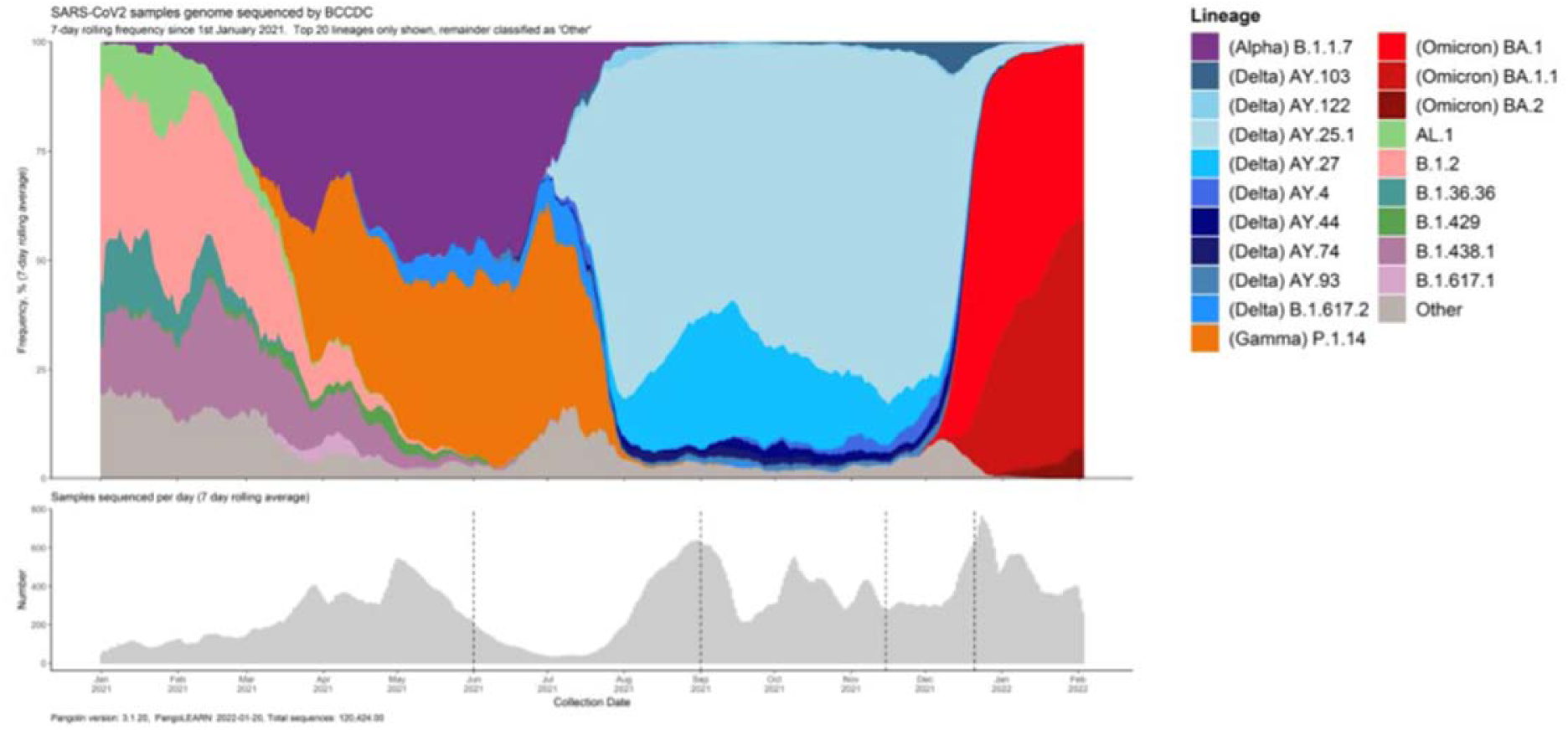
Twenty most prevalent SARS-CoV-2 variant of concern lineages in British Columbia from January 2021 to January 2022. The current study was performed during a time of Omicron variant predominance, from November19 2021 to January 8 2022. BCCDC: British Columbia Centre for Disease Control; SARS-CoV-2: severe acute respiratory syndrome coronavirus type 2

**Supplemental Figure 3.**
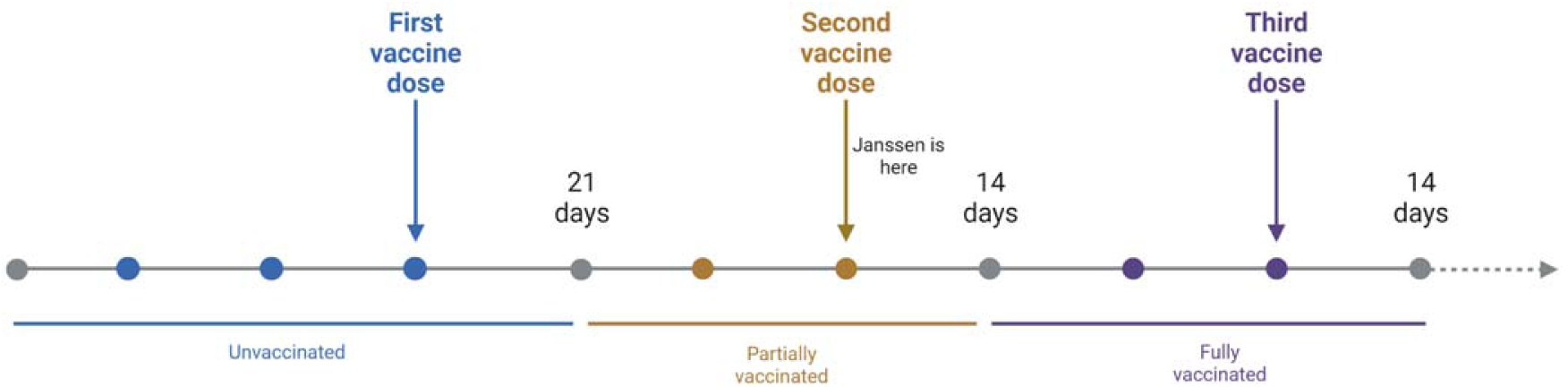
Vaccination status definitions. Vaccination status was defined based on the date of vaccine receipt relative to the date of the sample collection included for the study. For the Janssen vaccine only, fully vaccinated status was defined as having received one dose 14 days or more prior to sample collection. For all other vaccines, **Unvaccinated status** was defined as having received no SARS-CoV-2 vaccine, or having received a SARS-CoV-2 vaccine less than 21 days prior to the sample collection date. **Partially vaccinated** status was defined as having received the SARS-CoV-2 vaccine dose 1 greater or equal to 21 days prior to sample collection, but having received dose 2 less than 14 days prior to the sample collection. **Fully vaccinated** status was defined as greater or equal to 14 days since the receipt of dose 2, but having received dose 3 less than 14 days prior to the sample collection. SARS-CoV-2: severe acute respiratory syndrome coronavirus type 2

**Supplemental Figure 4.**
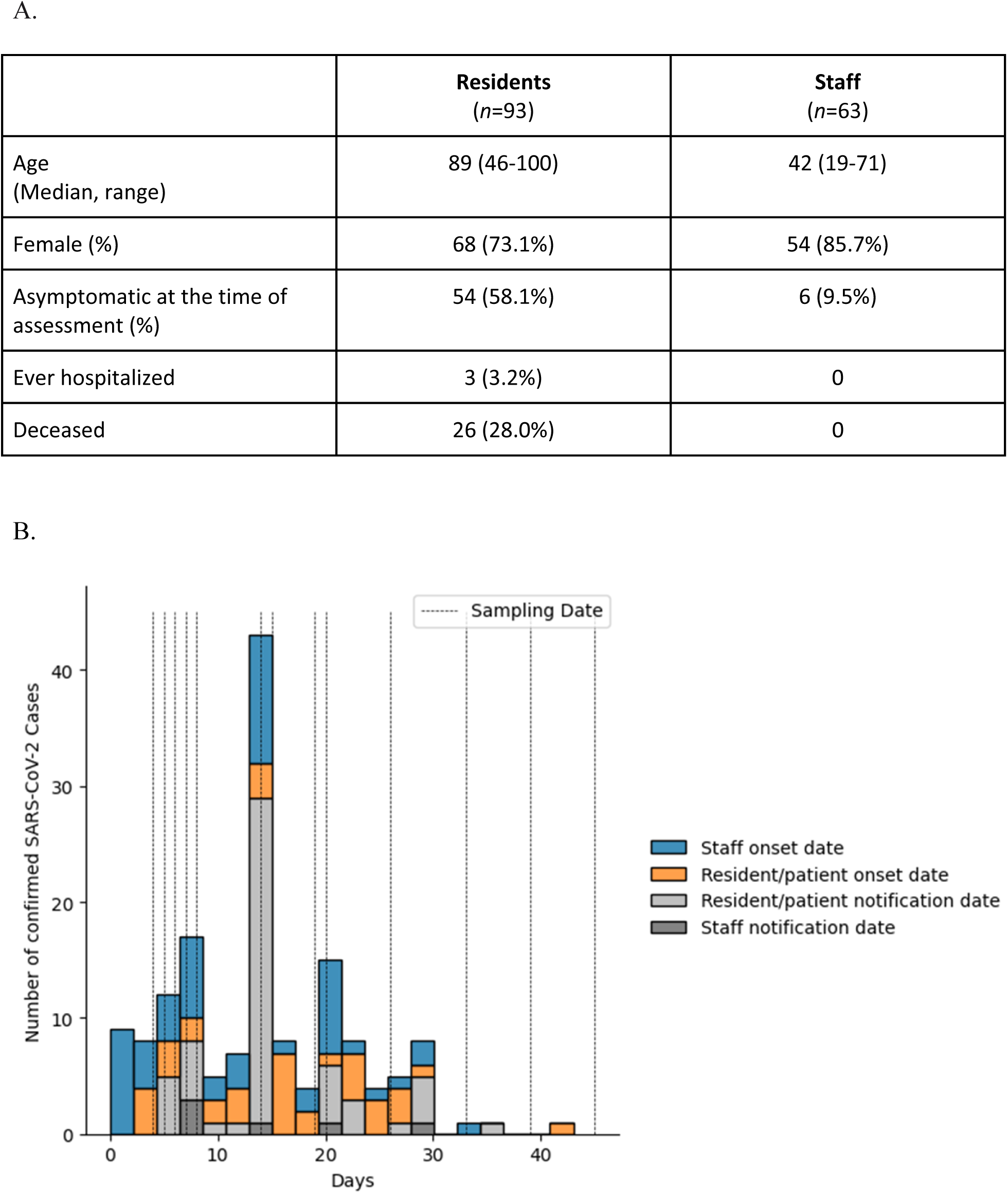
Case study epidemiological data (A) and epidemic curve (B) for the 156 infected individuals in the long term care facility outbreak.

**Supplemental Figure 5.**
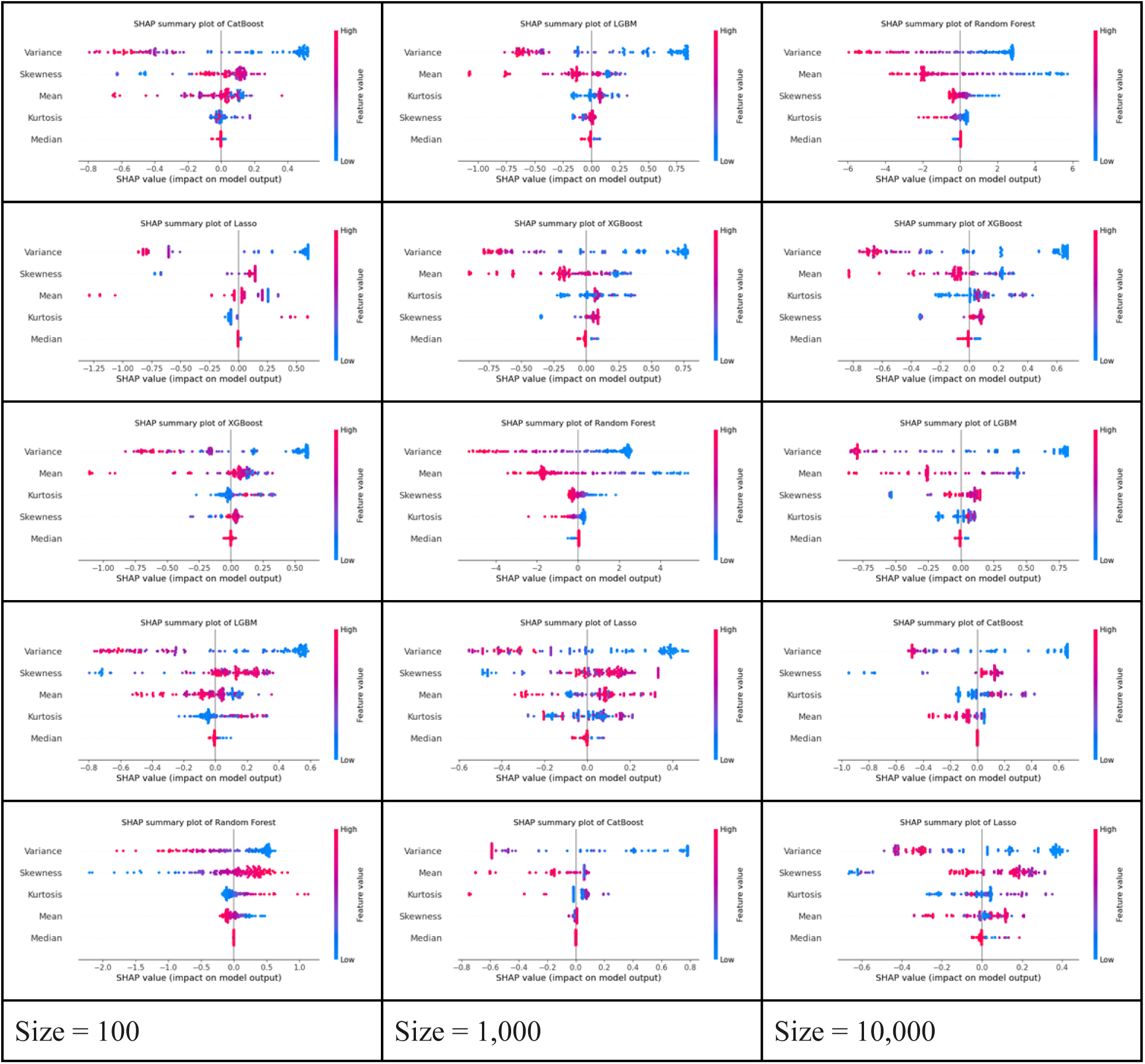
SHAP summary outputs explaining the machine learning output based on simulated cycle threshold (Ct) data. Results are presented stratified by three different population sizes: 100, 1,000 and 10,000 with each column in descending order of performance. Of the five features explored, the top ranking feature across all models was the variance of the Ct data. SHAP: SHapley Additive exPlanations; LGBM: Light Gradient Boosting Model; XGBM: eXtreme Gradient Boosting Model

## Notes

### Competing Interest Statement

The authors have declared no competing interest.

